# Development of a Diagnosis Grading System for Patients Undergoing Intrauterine Inseminations: A Machine-learning Perspective

**DOI:** 10.1101/2022.07.18.22277308

**Authors:** Changbo Jin, Jiaqi Zong, Shuya Xue

## Abstract

**Objective:** To develop an innovative, non-invasive and objective grading system for enhancing clinicians’ assessment of intrauterine insemination (IUI) patients.

**Design:** Patients who had undergone IUI treatments previously were divided into pregnant (N=4618) and non-pregnant(N=20974) groups. An evaluation index method was developed using collected clinical data from the two groups, particularly on indications of considerable differences between the two groups. The weight of each indicator was determined using random forest machine learning, and the indicators and patients’ conditions were classified using an entropy-based feature discretization technique. The indices for each indicator were further divided into five grades, from A to E, and given five points to one point, respectively. Effectiveness of the system was tested using the ten-fold cross-validation method.

**Setting:** Reproductive medicine center, Shanghai, China

**Patients:** Infertile couples who had undergone IUI treatment.

**Intervention:** None.

**Main Outcome Measures:** Weight of each indicator and grades of infertile patients.

**Results:** From the 25,592 medical records of infertile couples who had IUI, 4618 women were pregnant subsequently, with a mean age of 28.69±3.34 years. From the collected records, 18 indicators (e.g., body mass indices [BMI], endometrial thicknesses, couples’ ages, IUI cycle days, and semen situations) were selected to construct our diagnosis scoring system. Among the 18 indicators, BMI (weight, 12.49%), endometrial thickness (11.99%), female age (11.88%), semen density (10.41%), semen volume (8.92%), cycle day (7.38%) and male age (6.96%) were closely related to the pregnancy rates. Among patients with the final scores for > 75.29 individually, the pregnancy rates for them was > 56.35%. The system’s stability was 95.1% (95%CI,94.5%-95.7%) according to cross-validation data.

**Conclusion:** This quick and objective machine learning-based approach can be used to simplify and enhance the decision-making process among clinicians, especially to advise and to select patients for better IUI outcomes.

## Introduction

The incidence rates of infertility have been increasing around the world, especially in developed countries (1). According to statistics, 8-12% of childbearing couples in the world are affected by infertility(2). Consequently, various procedures have been developed to resolve the infertility problem, e.g., intrauterine insemination (IUI), in vitro fertilization (IVF) and intracytoplasmic sperm injection (ICSI). Compared to different procedures, advantages for IUI include minimal invasion, lower cost, less complications and higher compliances (3-5).

IUI is commonly performed in a natural female cycle, however, many factors such as hormone levels, female ages, ovarian reserve capacities, and body mass indices (BMI)(6, 7), can significantly affect the IUI outcomes. With impact from these variable data, clinicians had found it difficult to make quick and accurate decisions for efficient treatment plans for infertility patients.

To enhance efficacy for infertility patients, use of non-invasive prognostic factors has been recommended, e.g., ages, types of infertility, and durations of infertility (8-10).Artificial intelligence (AI), for example, including convolutional neural network (CNN), has already been widely used in enhancing diagnosis and treatment for various diseases (11-14); and random forest model and artificial neural network (ANN) have been used for improving accuracy in grading collected data (15-17). Indeed, the random forest, PLS and CNFE-SE models have been used in predicting clinical outcomes of IUI (18-20). However, these predictions can be improved by including a scoring system model for IUI. Therefore, this study was conducted by developing a non-invasive IUI scoring system to measure and to evaluate basic-line situations of patients who had undergone the IUI treatment, and to help clinicians in predicting pregnancy rates of these patients.

## Methods

### Data source and Collection

A retrospective study was performed by reviewing the clinical medical records of 31,933 IUI-treated patients at the Reproductive Medicine Center, from January 2017 to December 2021. The inclusion criteria for acceptance of the records were: (a) couples who were unable to conceive after 12 months of regular sexual intercourse without taking any contraceptive measures and were diagnosed as infertile. The diagnostic criteria for infertility were: unexplained infertility, mild endometriosis, cervical factors, and ovarian diseases. Criteria for male infertility were: oligospermia, asthenospermia, abnormal spermatozoa, semen liquefaction, and sexual dysfunction; (b) infertile patients who refused IVF and underwent at least one cycle of IUI; (c) infertile patients who had oviductography or laparoscopy showing at least one unclosed oviduct, or at least one side of the fallopian tube was not closed after salpingoplasty; (d) couples who received nature cycles and ovarian stimulation using FSH, clomiphene citrate (CC) or human menopausal gonadotropin (HMG). The collected data also included couples’ age, BMI, pregnancy history, semen routine, and other non-invasive examination items, excluding the results of sex hormone test, AMH, and other invasive tests.

### IUI indications and ovarian stimulation

Females who had IUI indications and had at least one of the following criteria would undergo ovarian stimulation: (a) with ovulation disorders, including polycystic ovary syndrome, hyperprolactinemia, ovarian hypofunction, and hypogonadotropic hypogonadism; (b) with normal ovulations but had not been pregnant for more than three months under the guidance of outpatient ovulation monitoring, or had not been pregnant after two natural cycles of artificial insemination, endometriosis, unexplained infertility.

All selected women underwent ovarian stimulation using clomiphene citrate (CC, Fertilan; Clomiphene Citrate Tablets; CODAL SYNTO Ltd. Cyprus) and HMG (Menotropins for Injection; Guangdong, Zhuhai, North Chuangye Road No. 38, Livzon Syntpharm Co., Ltd.; permission code of SFDA: H10940097). Patients were classified as follows: (a) Natural cycle group: No ovarian induction was performed; (b) Gonadotropins (Gn) group: given 75-150 IU of HMG or FSH daily by intramuscular injection starting from day 5 of the menstruation; (c) CC group: orally taking 50 or 100 mg of CC daily on cycle days 3 - 7 of the menstruation; (d) CC+Gn group: given 50 or 100 mg of CC daily on cycle days 3-7, followed by 75–150 IU of HMG or FSH daily via intramuscular injection.

### Measurement of follicles and endometrium

Transvaginal ultrasound was used to measure the presence and numbers of follicles. After adjusting for the frequency of the ultrasound probe, the probe was inserted through the posterior fornix of the vagina with a small amount of couplant to scan the patients’ uterus and other target areas in detail. The length and transverse diameter of each follicle were measured after the maximum section of the follicle was shown, and the average values were taken as the evaluation standards of the size for each follicle. The follicles with an average value ≥18mm each were considered as the dominant follicles in this cycle.

In order to measure thickness of the endometrium, sonogram of the sagittal plane of the uterus was taken and the maximum distance between the junction of the endometrium and the myometrium from both sides was measured. Measurements of endometrial thickness were performed by experienced sonographers with mm as the measurement unit. Each measurement was accurate to 0.01mm.

Before artificial inseminations, the patients were also routinely examined using transvaginal ultrasound to determine whether their follicles were released or not. If the follicles were not released, the patients were re-examined on the following days until follicles were released. Otherwise, these patients would not progress further. For patients who had released follicles, each patient was given progesterone 10 mg both in the mornings and evenings for 14 days. If menstruation did not come, the drug treatment would stop until the next cycle for artificial insemination. If menstruation came, luteal support would be provided to stop the cycle.

### Semen collection and insemination

Semen samples were collected from husbands/donors by masturbations and used as soon as possible for insemination. For husbands who were known to have testicular azoospermia and/or have family histories/genetic diseases for infertility, donor sperms were used instead.

In preparation for artificial insemination, female patients emptied the bladder, and positioned the uterus in the forward flexion and lithotomy situation. Normal saline was used to scrub the vulvas, vaginas, and external cervices. Washed semen samples were acknowledged by the doctors, nurses, and the patients before their use. For each patient, an insemination tube was used to siphon-up 0.5-1ml of a washed semen sample which was followed by taking up 0.1ml of air. The insemination tube was inserted into the uterine cavity in the direction of the uterus. The sample was slowly released into the uterine cavity at 1-2 cm outside the internal tube of the cervix. At about 1 minute after the release, the tube was pulled out and the patient was positioned with raised hip while in bed for at least 30 minutes.

### Constructing the IUI scoring system

Figure 1 shows the flow chart of establishing the system for this study. First, the collected data from the 18 indicators of each patient were reviewed to identify abnormalities and these abnormal data were eliminated. The reviewed data from all patients were then screened by the single-factor analysis of variance, and the weight of each index was calculated by a random forest classifier. The calculated weights were sorted from high to low and used as the core index of this scoring system.

**Figure 1.**
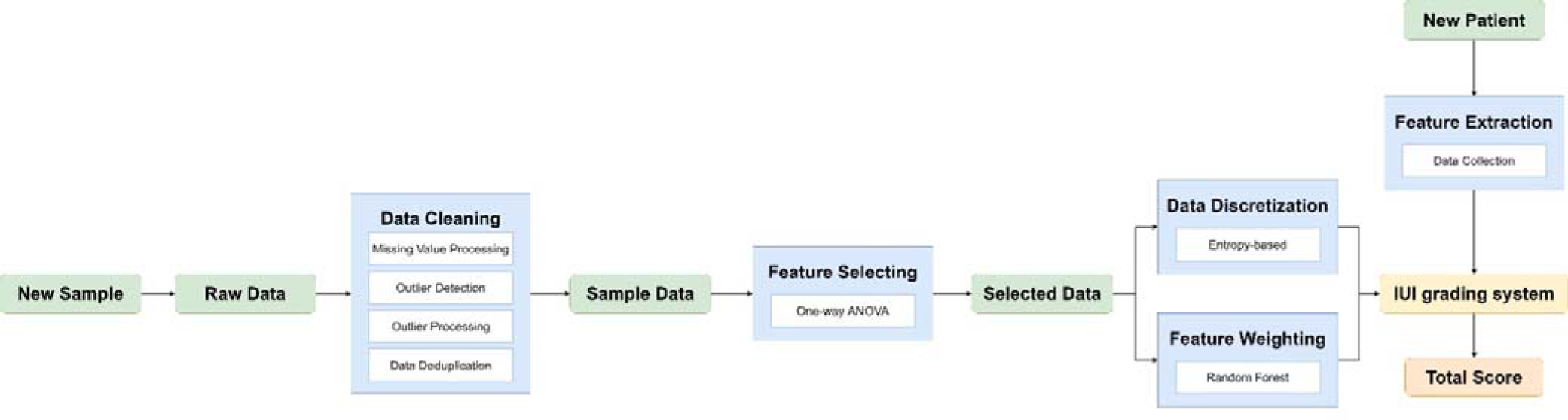
Flowchart of the IUI Dynamic Grading System

The collected data were also evaluated using a method based on information entropy to discretize the non-category features. This approach enhanced our effort to calibrate and to score the index values in different regions, and to obtain the total score for each patient. When all patients were scored according to the previously created scoring system,a ranking of total points for each patient were determined. The entropy segmentation method was used to divide the total score into five parts which were graded according to the pregnancy rates. Finally, a ten-fold cross-validation method was used to evaluate the classifier which would indicate that the scoring system had high reliability and could be used in the clinical process.

### Data review and processing

The work of data review and processing included identification of missing values, abnormal values, and data deduplications. In the missing value category, since only a small number from the large amount of collected data belonged to missing values, their deletions had little impact on the overall data. Consequently, attention was focused on outlier data. For this effort, the box chart which was not affected by outliers and did not require the data to obey normal distribution was prepared. The chart accurately and stably described the distribution of discrete data; therefore, they were conducive to use for data cleaning. The quartile distance (IQR) of the box chart was used to detect outliers, with the stipulation that the points that exceeded the upper quartile IQR distance or the lower quartile-1.5 times IQR distance were outliers. Subsequently, outliers with the maximum and minimum values were replaced by the next largest or smallest data within the normal values, respectively. After dealing with the missing values and outliers, 25592 pieces of data were available for further analyses.

### The calculation of random forest based on the index weight

The random forest classifier was built randomly and it contained multiple decision trees. Its output category was determined by the number of output types for each tree. The approach was suitable for dealing with high-dimensional data, for identifying important features and for calculation of indicator weight. In order to strike a balance between difficulties of training and effects of the model, the number of trees in the forest was set to be 100. In order to avoid the problem of data waste, the out-of-bag samples were used to consider the quality of the model, and scores outside the bag would reflect generalization ability of the model. After dividing the training test set according to 4:1 ratio, accuracy of the random forest classifier on the test set was 0.82 which indicated high accuracy. At the same time, contributions from each feature to each tree in the random forest were evaluated by taking the average value, comparing contributions between the features, characterizing importance of the 18 indexes in the classification model, and arranging them in the order from high to low.

### Feature discretization based on information entropy

The features based on information entropy were discretized to facilitate the scoring system for quantification of each index. The data discretization technique was used to group the continuous data of the 18 indicators into a discretized interval. The commonly used data discretization methods included equal width grouping, equal frequency grouping, etc. However, equal width grouping was more sensitive to discrete values and attribute values were unevenly distributed to each interval which could easily lead to data tilt. Although equal frequency grouping could avoid the shortcomings of equal width grouping, the same elements were divided into different groups, thus affecting the discrete results. Therefore, the data discretization method based on information entropy was chosen. The basic idea was to use the size of entropy to express the purity of the divided data set; the smaller the entropy, the higher the data purity, indicating the greater availability of discrete data. This approach was considered to be a better discretization method. The partition method was used to divide the data set into two parts, to calculate the sum of the entropy of the two parts, to divide them where the entropy was the least, and to repeat this step for the part with the most significant entropy until the numbers of data sets were reached that met the needs of users. After discretization, the interval of continuous features was collected, graded and scored based on each interval according to the clinical significance, to obtained the IUI scoring system.

### Performance evaluation of classifier based on ten-fold cross-validation

In order to reduce the chance caused by the single partition of the training set and test set, the existing data set was used to carry out multiple partitions, that is, through cross-validation to avoid selecting accidental hyper-parameters and models that did not have generalization ability because of particular partition. A ten-fold cross-validation divided all the data into ten parts, each as a verification set and the others as a training set for training and verification, and the average classification performance of 10 models was used to measure quality of the hyperparameters.

### Statistical Analysis

In order to test whether the factors played roles as a whole and/or influenced interactions between the factors, one-way ANOVA were conducted. Then, characteristics that played significant roles in pregnancy outcome were selected and they were used to construct a scoring system. The Python software (version: 3.7.4) was used to perform data pre-processing, random forest classifier weight calculations, feature discretization based on information entropy, and ten cross-validations. The Chi-squared (*χ*^2^) tests or Analysis of Variance (ANOVA) were conducted to compare indicators between groups. The P < 0.05 were considered as statistically significant when selecting the index in the scoring system.

## Results

### Key Indicators Selected to Construct the System

The study included 25592 couples who were treated with IUI and were separated into two groups based on whether or not they displayed typical pregnancy features: 4618 were in the pregnant group (mean [SD]: 28.69 [3.34] years), and 20974 in the non-pregnant group (29.35 [3.48] years). Detailed patient characteristics between two groups, including the age of both partners, BMI, semen parameters, infertility type, etc., are shown in Supplement Table 1. There were significant differences in the 18 indicators between the two groups: the pregnant group had significantly higher BMI (22.03[2.98] kg/m^2^ vs 21.83[2.89] kg/m^2^; P < 0.01), cycle days (14.29[2.48] days vs 14.13[2.54] days; P < 0.01) and greater endometrial thickness (9.52 [2.00] mm vs 9.36[2.08] mm; P < 0.01), and lower female age (28.69 [3.34] years vs 29.35 [3.48] years; P < 0.01), and fewer times of spontaneous abortions (0.10[0.40] vs 0.19[0.39], P < 0.05. There was no significant difference between the two groups in cycle numbers, times of delivery, and menarche ages. Consequently, our indicator system included 18 indicators: BMI, male and female age, endometrial thickness, six semen-related indexes, etc.

### Discretization Results of Indicators

Each of the 18 indicator indexes were divided into five grades using the entropy-based feature discretization method (Figure 2): A, B, C, D, and E, and were given scores of five, four, three, two, and one points, respectively. The scores represented the different abnormal levels of each indicator. The total score represented the sum of the weight-percentage corresponding to each feature score. The lower the scores, the worse the pregnancy outcomes.

**Figure 2.**
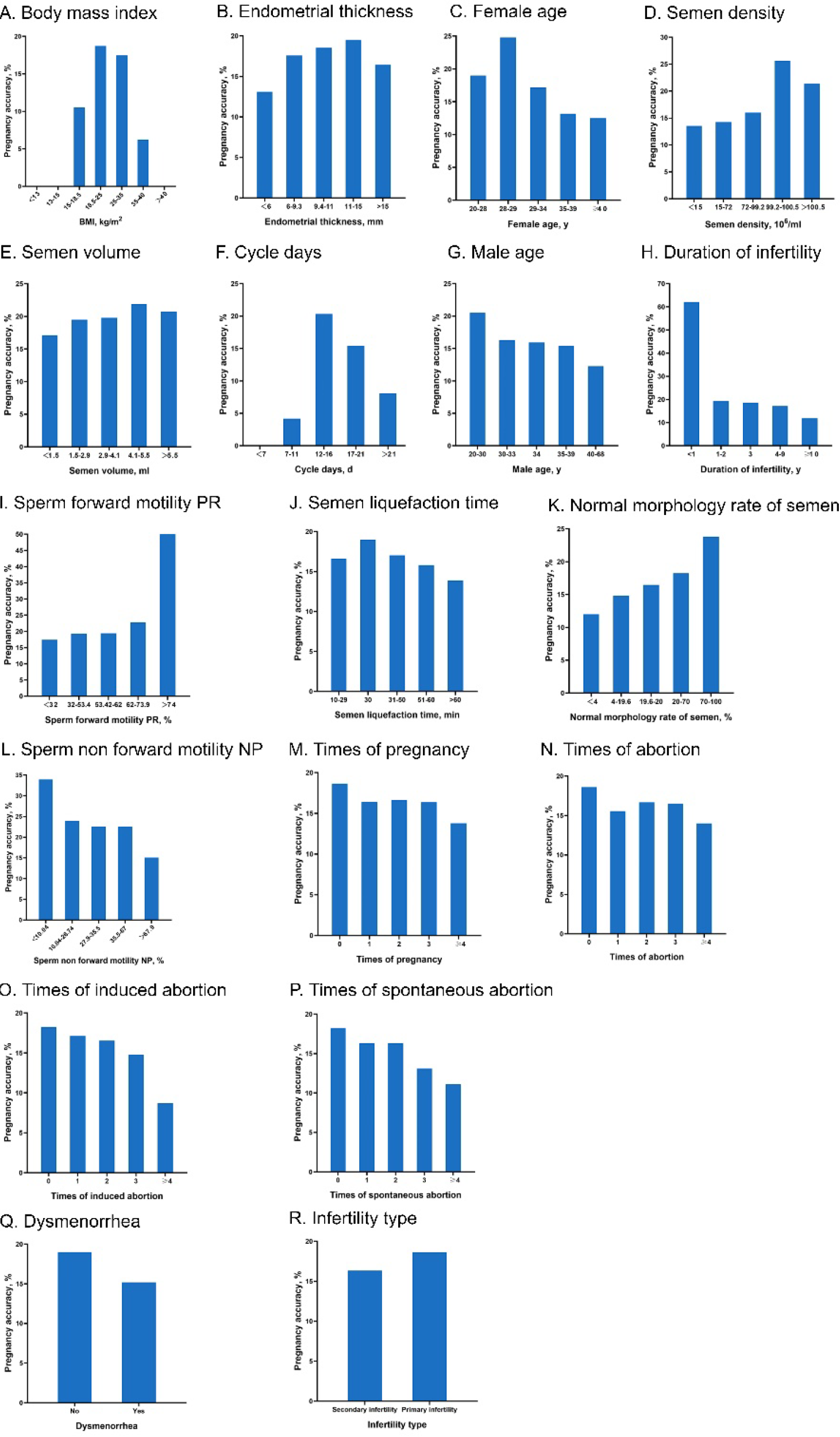
Interval Division Results of 18 Indicators

### Weight of Each Indicator

The random forest method used 80% of the data as the training set and 20% as the test set to assign weight to the 18 indicators. The weight of each index and the number of different categories of patients were distributed as shown in Table 1. Compared with other indexes, BMI (weight, 12.49%), endometrial thickness (11.99%), female age (11.88%), semen density (10.41%), semen volume (8.92%), cycle days (7.38%), male age (6.96%) had stronger correlation with the pregnancy rates.

**Table 1.** Grading and Weighting Results of 18 Indicators

| Indicators | Weight, % | Interval | Category | Score | Total sample | Pregnancy |  |
| --- | --- | --- | --- | --- | --- | --- | --- |
|  |  |  |  |  |  | Sample | Rate, % |
| BMI, kg/m<sup>2</sup> | 12.49 | < 13 | E | 1 | 0 | 0 | 0 |
|  |  | 13-15 | E | 1 | 0 | 0 | 0 |
|  |  | 15-18.5 | C | 3 | 2958 | 311 | 10.51 |
|  |  | 18.5-25 | A | 5 | 18714 | 3502 | 18.71 |
|  |  | 25-35 | B | 4 | 3888 | 679 | 17.46 |
|  |  | 35-40 | D | 2 | 32 | 2 | 6.25 |
|  |  | > 40 | E | 1 | 0 | 0 | 0 |
| Endometrial thickness, mm | 11.99 | < 6 | E | 1 | 954 | 125 | 13.10 |
|  |  | 6-9.3 | C | 3 | 10749 | 1890 | 17.58 |
|  |  | 9.4-11 | B | 4 | 9222 | 1708 | 18.52 |
|  |  | 11-15 | A | 5 | 4833 | 942 | 19.49 |
|  |  | >15 | D | 2 | 304 | 50 | 16.45 |
| Female age, y | 11.88 | 20-28 | B | 4 | 11364 | 2157 | 18.98 |
|  |  | 28-29 | A | 5 | 1319 | 327 | 24.79 |
|  |  | 29-34 | C | 3 | 10947 | 1876 | 17.14 |
|  |  | 35-39 | D | 2 | 1922 | 253 | 13.16 |
|  |  | ≥40 | E | 1 | 40 | 5 | 12.50 |
| Semen density, 10<sup>6</sup>/ml | 10.41 | < 15 | E | 1 | 2975 | 402 | 13.51 |
|  |  | 15-72 | D | 2 | 10583 | 1505 | 14.22 |
|  |  | 72-99.2 | C | 3 | 3795 | 608 | 16.02 |
|  |  | 99.2-100.5 | A | 5 | 8089 | 2071 | 25.60 |
|  |  | > 100.5 | B | 4 | 150 | 32 | 21.33 |
| Semen volume, ml | 8.92 | < 1.5 | E | 1 | 16782 | 2870 | 17.10 |
|  |  | 1.5-2.9 | D | 2 | 580 | 113 | 19.48 |
|  |  | 2.9-4.1 | C | 3 | 2865 | 567 | 19.79 |
|  |  | 4.1-5.5 | A | 5 | 4010 | 877 | 21.87 |
|  |  | > 5.5 | B | 4 | 1352 | 280 | 20.71 |
| Cycle days, d | 7.38 | < 7 | E | 1 | 58 | 0 | 0.00 |
|  |  | 7-11 | D | 2 | 2237 | 93 | 4.16 |
|  |  | 12-16 | A | 5 | 19622 | 3989 | 20.33 |
|  |  | 17-21 | B | 4 | 3266 | 503 | 15.40 |
|  |  | > 21 | C | 3 | 409 | 33 | 8.07 |
| Male age, y | 6.96 | 20-30 | A | 5 | 12433 | 2551 | 20.52 |
|  |  | 30-33 | B | 4 | 6886 | 1122 | 16.29 |
|  |  | 34 | C | 3 | 1603 | 255 | 15.91 |
|  |  | 35-39 | D | 2 | 3740 | 576 | 15.40 |
|  |  | 40-68 | E | 1 | 930 | 114 | 12.26 |
| Duration of infertility, y | 6.32 | <1 | A | 5 | 29 | 18 | 62.07 |
|  |  | 1-2 | B | 4 | 8789 | 1695 | 19.29 |
|  |  | 3 | C | 3 | 5323 | 988 | 18.56 |
|  |  | 4-9 | D | 2 | 10575 | 1813 | 17.14 |
|  |  | ≥10 | E | 1 | 876 | 104 | 11.87 |
| Sperm forward motility PR, % | 5.05 | < 32 | E | 1 | 16635 | 2898 | 17.42 |
|  |  | 32-53.4 | D | 2 | 4705 | 910 | 19.34 |
|  |  | 53.42-62 | C | 3 | 4070 | 788 | 19.36 |
|  |  | 62-73.9 | B | 4 | 180 | 41 | 22.78 |
|  |  | > 74 | A | 5 | 2 | 1 | 50.00 |
| Semen liquefaction time, min | 4.96 | 10-29 | C | 3 | 9850 | 1637 | 16.62 |
|  |  | 30 | A | 5 | 13417 | 2547 | 18.98 |
|  |  | 31-50 | B | 4 | 1481 | 252 | 17.02 |
|  |  | 51-60 | D | 2 | 772 | 122 | 15.80 |
|  |  | >60 | E | 1 | 72 | 10 | 13.89 |
| Normal morphology rate of semen, % | 4.90 | < 4 | E | 1 | 392 | 47 | 11.99 |
|  |  | 4-19.6 | D | 2 | 1203 | 178 | 14.80 |
|  |  | 19.6-20 | C | 3 | 152 | 25 | 16.45 |
|  |  | 20-70 | B | 4 | 23530 | 4293 | 18.24 |
|  |  | 70-100 | A | 5 | 315 | 75 | 23.81 |
| Dysmenorrhea | 2.18 | No | A | 5 | 19255 | 3656 | 18.99 |
|  |  | Yes | E | 1 | 6337 | 962 | 15.18 |
| Sperm non-forward motility NP, % | 1.45 | < 10.04 | A | 5 | 56 | 19 | 33.93 |
|  |  | 10.04-26.74 | B | 4 | 848 | 203 | 23.94 |
|  |  | 27.9-35.5 | C | 3 | 3658 | 825 | 22.55 |
|  |  | 35.5-67 | D | 2 | 5345 | 1204 | 22.53 |
|  |  | > 67.9 | E | 1 | 15685 | 2367 | 15.09 |
| Times of pregnancy | 1.36 | 0 | A | 5 | 18892 | 3521 | 18.64 |
|  |  | 1 | B | 4 | 4619 | 758 | 16.41 |
|  |  | 2 | C | 3 | 1365 | 227 | 16.63 |
|  |  | 3 | D | 2 | 513 | 84 | 16.37 |
|  |  | ≥4 | E | 1 | 203 | 28 | 13.79 |
| Times of abortion | 1.25 | 0 | A | 5 | 19828 | 3686 | 18.59 |
|  |  | 1 | B | 4 | 4172 | 649 | 15.56 |
|  |  | 2 | C | 3 | 1115 | 186 | 16.68 |
|  |  | 3 | D | 2 | 321 | 53 | 16.51 |
|  |  | ≥4 | E | 1 | 100 | 14 | 14.00 |
| Infertility type | 1.00 | Secondary infertility | E | 1 | 6199 | 1013 | 16.34 |
|  |  | Primary infertility | A | 5 | 19393 | 3605 | 18.59 |
| Times of induced abortion | 0.83 | 0 | A | 5 | 22614 | 4130 | 18.26 |
|  |  | 1 | B | 4 | 2342 | 401 | 17.12 |
|  |  | 2 | C | 3 | 435 | 72 | 16.55 |
|  |  | 3 | D | 2 | 88 | 13 | 14.77 |
|  |  | ≥4 | E | 1 | 23 | 2 | 8.70 |
| Times of spontaneous abortion | 0.69 | 0 | A | 5 | 23537 | 4287 | 18.21 |
|  |  | 1 | B | 4 | 1635 | 267 | 16.33 |
|  |  | 2 | C | 3 | 300 | 49 | 16.33 |
|  |  | 3 | D | 2 | 84 | 11 | 13.10 |
|  |  | ≥4 | E | 1 | 36 | 4 | 11.11 |

### A New Dynamic Diagnosis Grading System for Infertility

Each index score was weighed and each patient’s condition was determined by the total score after our calculations. The correlations between pregnancy rates and the final scores are shown in Supplement table 2: the higher the total scores, the better the pregnancy rates. According to our determinations, patients with a total score below 27.08 were classified as grade E, and with > 75.29 the pregnancy rate was at least 56.35%. The results from our ten-fold cross-validation are shown in Figure 3. The stability of the system was 95.1% (95%CI,94.5%-95.7%).

**Figure 3.**
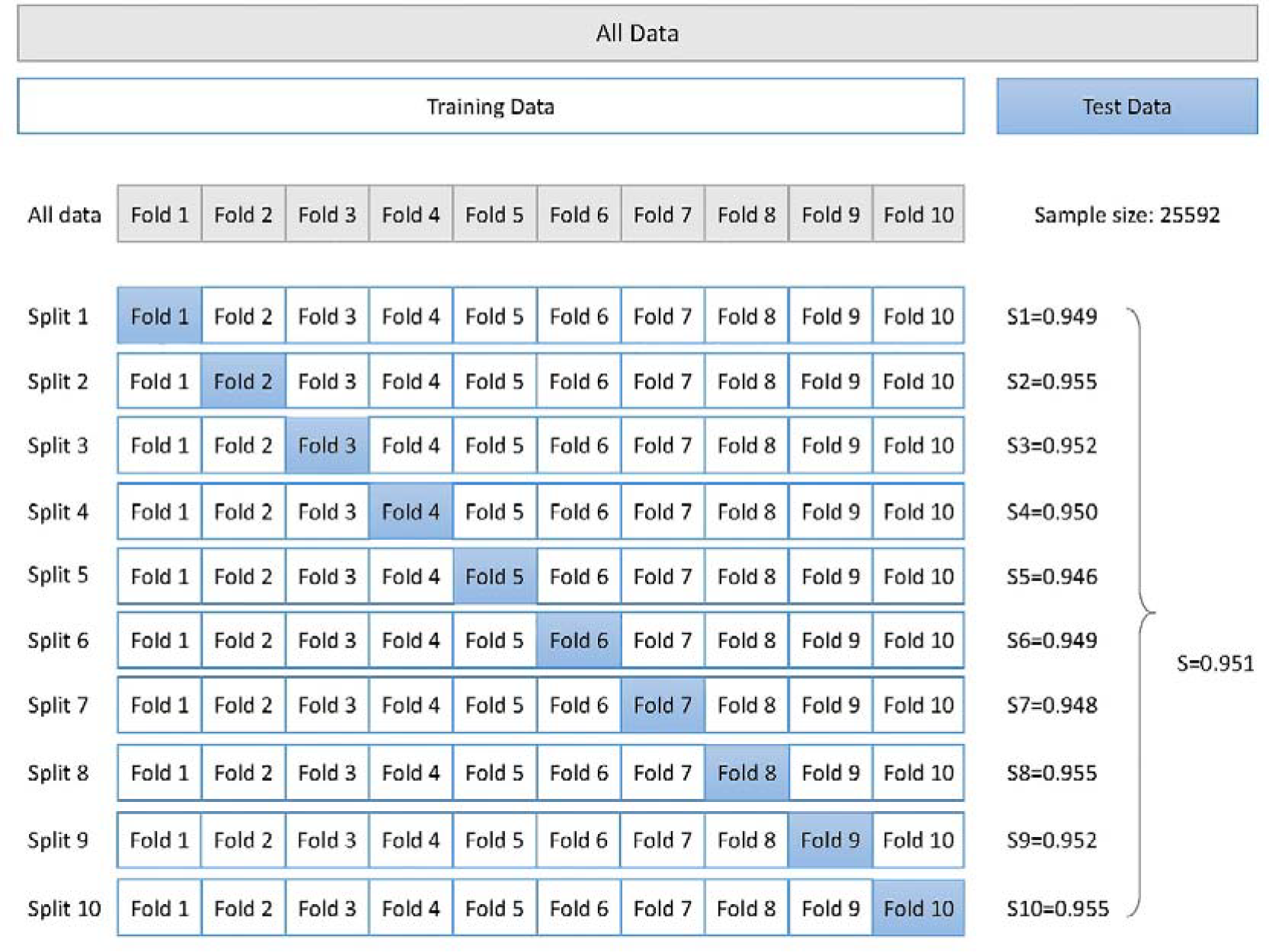
Ten-fold Cross Validation

### Association of Indicators with Pregnancy Rates

According to the changes in pregnancy rates and index values (Table 1), our grading system showed certain unique characteristics. First, the pregnancy rates decreased significantly when the BMI was less than 18.5kg/m^2^ or greater than 35 kg/m^2^. Secondly, when the endometrial thickness was between 11-15mm, the pregnancy rates were the highest. Third, when the cycle days were between 12-16 days, the pregnancy rates were the highest, and beyond this range, the pregnancy rates declined. Fourth, when women were younger than 29 years old, ages were positively correlated with the pregnancy rates. The pregnancy rates of women after the age of 29 showed a downward trend, especially after 34. Fifth, higher male ages were associated with lower pregnancy rates, and the pregnancy rates decreased significantly after age 30. Sixth, the pregnancy rates showed a downward trend when the durations of infertility were longer. Seventh, higher semen volumes, semen densities, sperm forward motility rates, but lower sperm deformity rates were associated with the higher pregnancy rates. Eighth, with increased abortion times, whether natural or induced abortions, the pregnancy rates decreased. Ninth, patients without dysmenorrhea had higher pregnancy rates than patients with dysmenorrhea. Tenth, patients with secondary infertility had higher pregnancy rates than patients with primary infertility.

## Discussion

In order to significantly enhance clinicians’ capacity to evaluate patients for IUI and to predict outcomes, a new and non-invasive IUI scoring system was developed by us. The system was generated based on collected data from 18 indicators (e.g., body mass indices [BMI], endometrial thicknesses, couples’ ages, IUI cycle days, and semen situations) from > 25,500 infertile couples, and on analyses using random forest algorithm and entropy-based feature discretization algorithm. To our knowledge, this is the first time an artificial intelligence method was used to construct an IUI grading system for patient assessment. Several major conclusions were derived from our single investigation: 1) patients who were too thin or obese had reduced pregnancy rates; 2) increased infertility durations led to lower pregnancy rates; 3) higher semen volume, semen density, sperm forward motility rate, but lower % deformed sperms were associated with higher pregnancy rates; 4) endometrial thicknesses were positively correlated with pregnancy rates when the endometrial thickness was less than 15 mm; 4) when the cycle days were between 12-16 days, the pregnancy rate was the highest, and beyond this range, the pregnancy rates declined; 5) men’s age and number of times of abortion were adversely connected with lower pregnancy rates; 6) the pregnancy rates of patients with primary infertility were higher than that of patients with secondary infertility; 7) Pregnancy rates of patients without dysmenorrhea were higher than those with dysmenorrhea.

Our conclusions have support from several investigations. Women who were younger than 40 years old, with a BMI less than 25 were higher pregnancy after IUI treatment (10). Abnormal sperm morphology and reduced sperm motility were prognostic factors for lower pregnancy rate of IUI (18-20). Pregnancies were increased with increased thickness in endometrium (21, 22). In addition, infertility patients with gynecological diseases such as adenomyosis and endometriosis had reduced pregnancy rates with assisted reproductive technology compared to healthy but infertile patients (23-26).

Based on our analyses using data from 18 clinical indicators, an IUI scoring system for predicting IUI outcomes was developed. The system is simple and dynamic because more indicators can be added, after their evaluation and validation. At this stage, our investigation indicates that patients whose final score was > 75.29 individually, the pregnancy rate for the group was > 56.35%. The system’s stability was 95.1% (95%CI, 94.5%-95.7%) according to cross-validation data. Such information should be critically useful for clinicians to evaluate patients and to predict pregnancy outcomes from IUI treatments with more confidence.

### Limitations

There are a few drawbacks to be noted. First, because of the small sample size of individual indicators in specific ranges, an abnormality between the interval of individual indicators and the pregnancy rate may exist. Furthermore, the data for this study came solely from a single hospital’s medical records. Even if the sample size was large enough, regional and population constraints may exist. For all of these limitations, the findings need to be interpreted with caution before being replicated in other multi-regional studies with larger sample size.

## Conclusions

The new non-invasive IUI grading system in this study can be used to assist clinicians in making quick and accurate preliminary assessments of IUI patients’ health. Aside from fully comprehending the severity of the patient’s conditions, this system can also be used to predict the pregnancy rate of patients who have undergone IUI treatment, with the treatment plan specifically selected.

## Data Availability

All data produced in the present study are available upon reasonable request to the authors

