## supplement for "Development of a Diagnosis Grading System for Patients Undergoing Intrauterine Inseminations: A Machine-learning Perspective": Supplemental tables.docx

**Supplemental table 1. Comparisons between the pregnant and the non-pregnant groups**

| **Variables** | **Pregnancy (+)**  **(N=4618)** | **Non-pregnancy (-)**  **(N=20974)** | **P-value** |
| --- | --- | --- | --- |
| BMI (kg/m^2^) | 22.03±2.98 | 21.83±2.89 | 2.19e-4 |
| Endometrial thickness (mm) | 9.52±2.00 | 9.36±2.08 | 1.99e-6 |
| Semen density (10^6^/ml) | 55.80±50.03 | 47.79±49.08 | <2e-16 |
| Semen volume (ml) | 3.09±1.52 | 1.87±1.40 | <2e-16 |
| Cycle days (days) | 14.29±2.48 | 14.13±2.54 | 1e-5 |
| Male age (years) | 30.37±4.09 | 31.07±4.31 | <2e-16 |
| Female age (years) | 28.69±3.34 | 29.35±3.48 | <2e-16 |
| Duration of infertility (years) | 3.56±2.20 | 3.81±2.38 | 8.62e-10 |
| Sperm forward motility PR, % | 33.22±17.52 | 31.29±16.58 | 6.48e-14 |
| Semen liquefaction time (min) | 39.49±11.01 | 44.57±28.31 | <2e-16 |
| Normal morphology rate of semen (%) | 20.60±6.32 | 19.76±6.72 | 3.13e-5 |
| Cycle number | 1.85±1.00 | 1.86±1.01 | 0.843 |
| Sperm non forward motility NP (%) | 10.90±7.73 | 12.50±7.98 | 1.18e-9 |
| Times of pregnancy | 0.36±0.78 | 0.39±0.79 | 6.07e-5 |
| Times of abortion | 0.29±0.70 | 0.32±0.70 | 6.62e-5 |
| Times of induced abortion | 0.13±0.45 | 0.15±0.44 | 0.014 |
| Times of spontaneous abortion | 0.10±0.40 | 0.19±0.39 | 1.48e-5 |
| Times of delivery | 0.04±0.19 | 0.04±0.18 | 0.506 |
| Menarche age | 13.32±1.10 | 13.39±1.02 | 0.764 |
| Infertility type | | | |
| Primary | 3605(78.06%) | 15788(75.27%) | 6.17e-5 |
| Secondary | 1013(21.94%) | 5186(24.73%) |  |
| Dysmenorrhea | | | |
| Yes | 962(20.83%) | 5375(25.63%) | 8.20e-12 |
| No | 3656(79.17%) | 15599(74.37%) |  |

Data are mean ± SD or number (percentage). BMI: body mass indices

|  |  |  | **Pregnancy** |  |
| --- | --- | --- | --- | --- |
| **Categories** | **Total score** | **Total sample** | **Sample** | **Rate, %** |
| E | ＜27.08 | 13 | 1 | 7.69% |
| D | 27.08-43.82 | 4943 | 697 | 14.10% |
| C | 43.82-61.94 | 9879 | 1610 | 16.30% |
| B | 61.94-75.29 | 10631 | 2249 | 21.16% |
| A | ＞75.29 | 126 | 71 | 56.35% |

**Supplemental table 2. Pregnancy Rates and Total Scores for Patients**
